# Phenotypic effect of *GBA1* variants in individuals with and without Parkinson disease: the RAPSODI study

**DOI:** 10.1101/2023.07.25.23291637

**Authors:** M Toffoli, H Chohan, S Mullin, A Jesuthasan, S Yalkic, S Koletsi, E Menozzi, S Rahall, N Limbachiya, N Loefflad, A Higgins, J Bestwick, S Lucas-Del-Pozo, F Fierli, A Farbos, R Mezabrovschi, C Lee-Yin, A Schrag, D Moreno-Martinez, D Hughes, A Noyce, K Colclough, AR Jeffries, C Proukakis, AHV Schapira

## Abstract

**Background:** Variants in the *GBA1* gene cause the lysosomal storage disorder Gaucher disease (GD). They are also risk factors for Parkinson disease (PD), and modify the expression of the PD phenotype.

The penetrance of *GBA1* variants in PD is incomplete, and the ability to determine who amongst *GBA1* variant carriers are at higher risk of developing PD, would represent an advantage for prognostic and trial design purposes.

**Objectives:** To compare the motor and non-motor phenotype of *GBA1* carriers and non-carriers.

**Methods:** We present the cross-sectional results of the baseline assessment from the RAPSODI study, an online assessment tool for PD patients and *GBA1* variant carriers. The assessment includes clinically validated questionnaires, a tap-test, the University of Pennsyllvania Smell Identification Test and cognitive tests. Additional, homogeneous data from the PREDICT-PD cohort were included.

**Results:** A total of 379 participants completed all parts of the RAPSODI assessment (89 *GBA1*-negative controls, 169 *GBA1*-negative PD, 47 *GBA1*-positive PD, 47 non-affected *GBA1* carriers, 27 GD). Eightysix participants were were recruited through PREDICT-PD (43 non-affected *GBA1* carriers and 43 *GBA1*-negative controls). *GBA1*-positive PD showed worse performance in visual cognitive tasks and olfaction compared to *GBA1*-negative PD patients. No differences were detected between non-affected *GBA1* carriers carriers and *GBA1*-negative controls. No phenotypic differences were observed between any of the non-PD groups.

**Conclusions:** Our results support previous evidence that *GBA1*-positive PD has a specific phenotype with more severe non-motor symptoms. However, we did not reproduce previous findings of more frequent prodromal PD signs in non-affected *GBA1* carriers.

## INTRODUCTION

The *GBA1* gene encodes the lysosomal enzyme glucerebrosidase. Variants in *GBA1* are a risk factor for Parkinson disease (PD)(1), with a penetrance that is variable and ranges according to the severity of the variant(2).

The clinical phenotype of PD seems to be significantly worse in patients that carry *GBA1* variants compared to non-carriers, although how domains differ and to what extent are matters of debate(3–5). *GBA1* variant carriers have an earlier age of PD onset, with poorer overall cognitive function(3), more frequent non-motor symptoms, visual hallucinations and motor complications(6,7). Some data also suggests a higher prevalence of pre-clinical symptoms in healthy *GBA1* variants carriers compared to non-carriers(8–11), although this has not been replicated in independent cohorts(12).

Understanding the role of *GBA1* variants in determining phenotypic characteristics is important for prognostic purposes, and to guide the design of clinical trials.

Here, we report baseline data from the homogenous cohorts RAPSODI (rapsodistudy.com) (13) and PREDICT-PD (predictpd.com), online cohorts for remote assessment of motor and non-motor signs of parkinsonism. We compare characteristics of PD patients with and without *GBA1* variants, healthy *GBA1* carriers, Gaucher disease (GD) patients and controls. We hope to provide further insight into the phenotype-genotype correlation of *GBA1* variants in the pathogenesis of PD.

## METHODOLOGY

### Recruitment of participants

Participants were recruited through RAPSODI (rapsodistudy.com)(13). The study commenced active recruitment in January 2018 and participants are asked to repeat the assessment every year for up to 25 years. In this paper, we report data from the baseline (year 1) assessment. Participants were allowed to join the study if they were between the age of 18 and 90 and if they: had a diagnosis of GD, a diagnosis of PD, if they knew they carried a *GBA1* variant or if they were relatives of a PD patient, GD patient or *GBA1* variant carrier. Exclusion criteria were the presence of dementia or any other conditions known to cause parkinsonism. Upon enrollment, all participants were required to give informed consent to be included in the study. The work was approved by the London – Queen Square Research Ethics Committee (REC reference: 15/LO/1155).

### Assessment

A detailed description of the study design can be found in a previous publication(13). Participants were asked to complete the the REM Sleep Behavior Disorder Questionnaire (RBDsq)(14), the Unified Parkinson Disease Rating Scale part 2 (MDS-UPDRS2)(15) and the Hospital Anxiety and Depression Scale (HADS)(16). The RBDsq has been validated in the general population with a cut-off of 5. However, in this study a cut-off of 6 was used, as it is considered more appropriate for people with PD(17). Established cut-offs for the HADS scale (0-7 Normal, 8-10 Borderline and 11-21 Abnormal) were used for the sub-scores of depression and anxiety(16).

Additionally, participants were asked 3 questions about constipation: “Does opening your bowels require a lot of effort?”, “Do you suffer from hard stools?”, “Do you ever use laxatives?”. These had multiple choice answers “Yes”, “Sometimes” and “No”.

Cognitive Tests were delivered through the ‘CogTrack™’ platform(18), investigating different aspects of cognition, including pattern separation ability, simple reaction time, choice reaction time, digit vigilance, spatial working memory and numeric working memory. A summary of the tests and outcomes used can be found in table 1.

**Table 1.** Cognitive Tests.

| Test | Description of test | Outcome measure | Description of outcome |
| --- | --- | --- | --- |
| Pattern separation test | Participants are shown 20 pictures. After 10 minutes, they are re-presented with the images alongside 20 similarly looking images, to determine if they could detect the original ones. | DPICOACC | Percentage of correct answers recognising original pictures |
|  |  | DPICNACC | Percentage of correct answers recognising new pictures |
| Simple reaction time | measures the speed of participants to elicit a simple motor response to an expected stimulus, which occurred repeatedly but at unpredictable intervals. | SRT | Median reaction time (ms) |
| Choice reaction time | Participants are asked to monitor a screen for one of two possible stimuli, which occurred at unpredictable intervals. They must press a key dependent on which of the two stimuli was shown. | CRT | Median reaction time (ms) |
|  |  | CRTACC | Percentage of correct answer |
| Digit vigilance | Participants monitor a series of digits presented individually and rapidly in the centre of the screen and press the right arrow when the digit matches the target digit displayed constantly on the right of the screen. | VIGRT | Median reaction time (ms) |
|  |  | VIGACC | percentage of targets detected |
| Spatial working memory | Participants are shown three rows of 3 light bulbs, of which 4 of the bulbs were lit. They are then presented with the 3 rows of bulbs, each time with only one of them lit. They must respond as to whether the lit bulb is one of the original 4 that were lit up. | SPMRT | Median reaction time (ms) |
|  |  | SPMOACC | Percentage of correct 'yes' answers |
|  |  | SPMNACC | Percentage of correct 'no' answers |
| Numeric working memory | Participants are shown a series of 5 different digits, one digit at a time, and then a series of single digits to which they must answer as quickly as possible as to whether the digit was in the original series. | NWMRT | Median reaction time (ms) |
|  |  | NWMOACC | Percentage of correct 'yes' answers |
|  |  | NWMNACC | Percentage of correct 'no' answers |

The BRadykinesia Akinesia INcoordination (BRAIN) test(19,20) was used to evaluate hand dexterity and bradykinesia, in which participants were asked to press the “S” and “;” keys on their keyboard in succession as fast as they could. Each hand was assessed separately for 30 seconds and all participants were given a preceding 5 second practice trial before data was collected. The kinesia score (KS30), corresponding to the number of taps in 30 seconds, as well as the akinesia time (AT30), which was the mean dwell time on each key in milliseconds (msec) were calculated.

Olfactory function was measured using the University of Pennsylvania Smell Identification Test (UPSIT)(21). The cut-offs provided by the UPSIT manual identified different degrees of deficit: anosmia (0-18), severe microsmia (19-25), moderate microsmia (26-29 for males, 26-30 for females), mild microsmia (30-33 for males, 31-34 for females), and normosmia (34-40 for males, 35-40 for females).

### Collection of saliva samples and sequencing

Saliva samples were collected with the DNA OG-500 kit from DNA genotek, posted to participants upon completion of the online part of the assessment. Sequencing of the *GBA1* gene was carried out at the University of Exeter Sequencing Facility with a long read, nanopore technology method previously described(22). The *LRRK2* G2019S variant was genotyped with KBiosciences Competitive AlleleSpecific PCR SNP genotyping system by an external laboratory (LGC Genomics, Hoddesdon, Herts).

### PREDICT-PD

To seek further validation, additional non-affected *GBA1* carriers and age and sex matched *GBA1*-negative controls were included from the PREDICT-PD study. PREDICT-PD is a web-based cohort study to identify individuals at higher risk of PD (ref Noyce et al JNNP 2014). *GBA1* variants were identified through Sanger sequencing of exons 8-11, rather than full gene sequencing (Noyce et al, Movement Disorders 2017). Questions about constipation, RBDsq, HADS, UPSIT and tap-test were collected similarly to RAPSODI. For these, results show the combined data from the two cohorts. CogTrack testing was not available for PREDICT-PD and are thus only reported for the RAPSODI cohort.

### Statistical analysis

R version 4.2.2 was used for statistical analyses.

All outcome measures were compared between the 5 groups. Additional sub-analysis were carried out comparing carriers of risk, mild and severe *GBA1* variants(23).

ANOVA was used to assess differences in age, disease duration, age at diagnosis, years of education, with Tukey multiple comparison test for *post hoc* analysis.

Ordinal logistic regression was used to analyse questions about constipation, MDS-UPDRS2 (after dividing the values in equal deciles), anxiety and depression subscores of HADS, and UPSIT. Logistic regression was used to analyse outcomes of the RBDsq. Linear regression was used to assess differences in KS30, AT30, SRT, CRT, VIGRT, SPMRT, NWMRT. The cognitive scores for accuracy (DPICOACC, DPICNACC, CRTACC, VIGACC, SPMOACC, SPMNACC, NWMOACC, NWMNACC) represent proportions of correct answers, so they were analysed with quasibinomial regression. Age and sex were used as covariate in all analysis, and education was used as covariate in the cognitive tests. Outliers, defined as observations more than 3 standard deviations from the mean, were removed from the tap test and cognitive test scores.

## RESULTS

Anonymised participant-level data are reported as supplementary material.

### Size, demographics and genotype

Size and demographics of the cohort of participants that completed the whole assessment are reported in table 2. One participant had both GD and PD and was excluded from the analysis. Two PD participants were found to carry the *LRRK2* p.G2019S variant and were also excluded from the analysis. Not all participants completed all steps of the assessment, so numbers vary for each test.

**Table 2.** Demographics of participants that completed the every part of the assessment.

| Study | status | N | Age (mean ± SD) | PD duration in years (mean ± SD) | Age at onset of PD (mean ± SD) | Males (percentage) | Females (percentage) | Years of education (mean ± SD) |
| --- | --- | --- | --- | --- | --- | --- | --- | --- |
| *RAPSODI* | *GBA1*-negative controls | 89 | 55.9 ± 13.6 | NA | NA | 24 (27.0%) | 65 (73.0%) | 14.9 ± 13.6 |
| | *GBA1*-negative PD patients | 169 | 64.3 ± 8.7 | 3.1 ± 8.7 | 61.4 ± 8.7 | 89 (52.7%) | 80 (47.3%) | 14.5 ± 8.7 |
| | *GBA1*-positive PD patients | 47 | 60.1 ± 9.8 | 4.5 ± 9.8 | 58.0 ± 9.8 | 30 (63.8%) | 17 (36.2%) | 14.4 ± 9.8 |
| | GD Patients | 27 | 54.4 ± 15.2 | NA | NA | 16 (59.2%) | 11 (40.7%) | 13.8 ± 15.2 |
| | Non-affected *GBA1* carriers | 47 | 51.9 ± 15.5 | NA | NA | 18 (38.3%) | 29 (61.7%) | 14.6 ± 15.5 |
| PREDICT-PD | *GBA1*-negative controls | 43 | 67.5 ± 5.0 | NA | NA | 20 (46.5%) | 23 (53.5%) | NA |
| | Non-affected *GBA1* carriers | 43 | 67.5 ± 5.0 | NA | NA | 20 (46.5%) | 23 (53.5%) | NA |
| *Unified cohorts* | *GBA1*-negative controls | 132 | 59.7 ± 12.7 | NA | NA | 44 (33.3%) | 88 (66.7%) | NA |
| | *GBA1*-negative PD patients | 169 | 64.3 ± 8.7 | 3.1 ± 8.7 | 61.4 ± 8.7 | 89 (52.7%) | 80 (47.3%) | NA |
| | *GBA1*-positive PD patients | 47 | 60.1 ± 9.8 | 4.5 ± 9.8 | 58.0 ± 9.8 | 30 (63.8%) | 17 (36.2%) | NA |
| | GD Patients | 27 | 54.4 ± 15.2 | NA | NA | 16 (59.2%) | 11 (40.7%) | NA |
| | Non-affected *GBA1* carriers | 90 | 59.4 ± 14.0 | NA | NA | 38 (42.2%) | 52 (57.8%) | NA |

Age at recruitment for *GBA1*-negative PD patients was significantly higher than for *GBA1*-negative controls, GD patients and non-affected *GBA1* carriers (all p-values <0.01). No other significant differences in age at recruitment were observed. Sex was significantly different between the groups (p-value <0.001).

There were no significant differences in disease duration or age at diagnosis among the PD groups. Years of education were similar between the groups.

Genotypes of *GBA1* positive participants are reported in table 3 and in more details in supplementary table 1.

**Table 3.**
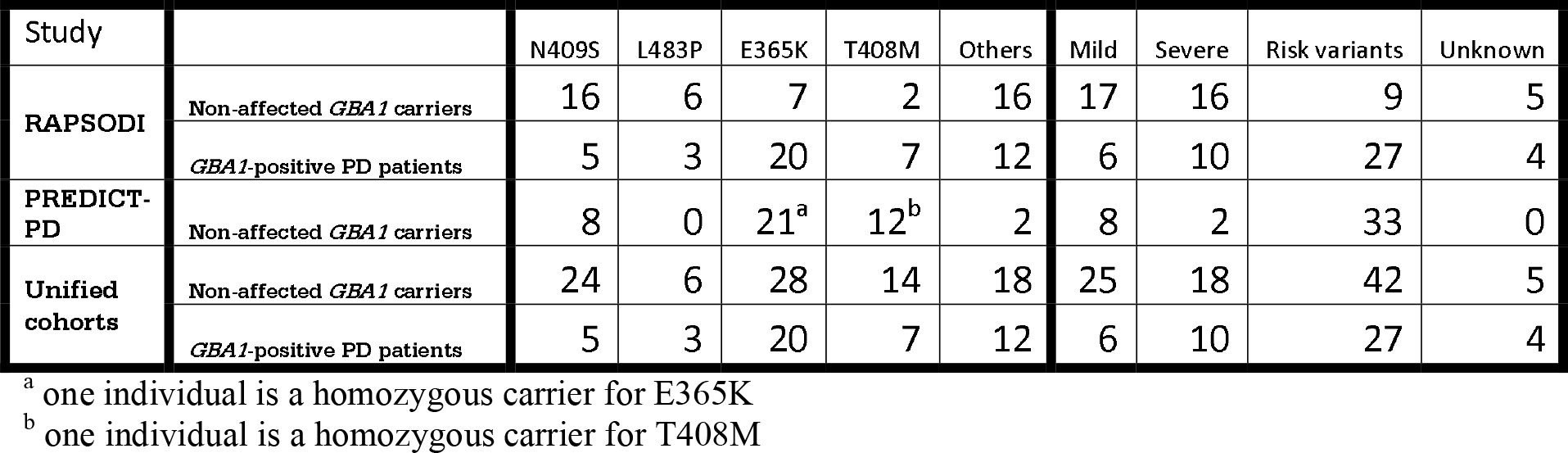
Genotype of *GBA1* variants carriers.

### Questionnaires and UPSIT

Questionnaire results are reported in Supplementary Table 2 and in Figure 1.

**Figure 1.**
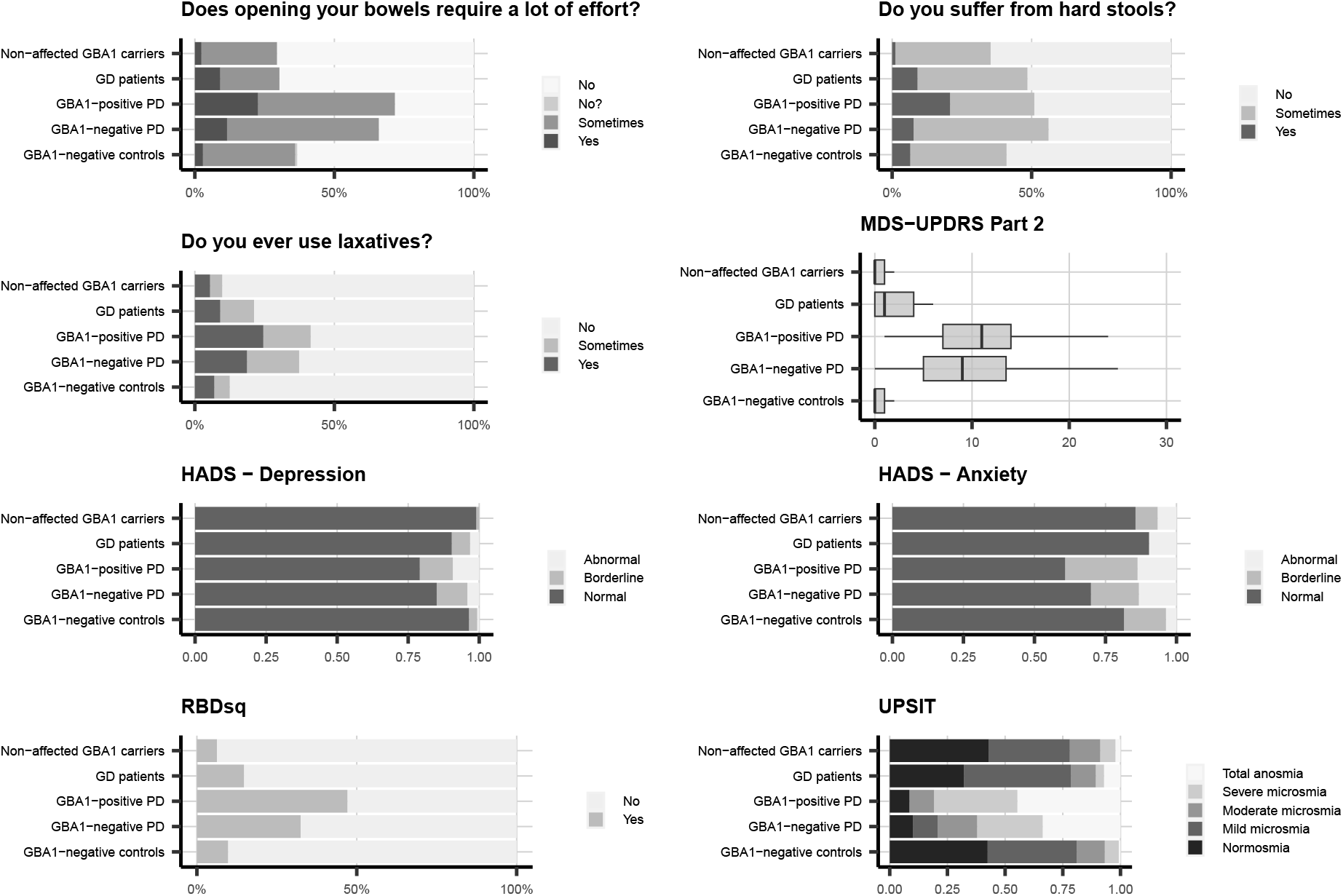
Clinical questionnaires and UPSIT For constipation, HADS, RBDsq and UPSIT scores, data are reported as percentage of participants per group. MDS-UPDRS 2 scores are reported as mean (central bar), 25^th^ and 75^th^percentiles (hinges) and the smallest value at most 1.5 * interquartile range of the hinge (whiskers). MDS-UPRS: Movement Disorder Society – Unified Parkinson Disease Rating Scale. HADS: Hospital Anxiety and Depression Scale. RBDsq: REM Sleep Behaviour Disorder screening questionnaire. UPSIT: University of Pennsylvania Smell Identification Test.

The two PD groups performed worse than all the non-PD groups in the questions about constipation (all p-values < 0.05), in the MDS-UPDRS2 (all p-values < 0.001), anxiety subscores of HADS (all p-values < 0.05), RBDsq (all p-values < 0.05), UPSIT (all p-values < 0.001).

The depression sub-score of HADS showed worse outcomes for the two PD groups compared to non-affected *GBA1* carriers and *GBA1*-negative controls (p-values all < 0.05), but no differences between the PD groups and GD patients.

*GBA1*-positive PD patients scored worse than *GBA1*-negative PD patients in UPSIT (p-value 0.015, OR 0.47, CI 0.25-0.86).

No differences were observed between any of the non-PD groups for any of the questionnaires or UPSIT.

No differences were observed between risk, mild and severe variant carriers among *GBA1*-positive PD and non-affected *GBA1*-carriers.

Results did not change when analysing the RAPSODI cohort separately.

### Tap test

Tap test results are reported in supplementary table 3 and in Figure 2.

**Figure 2.**
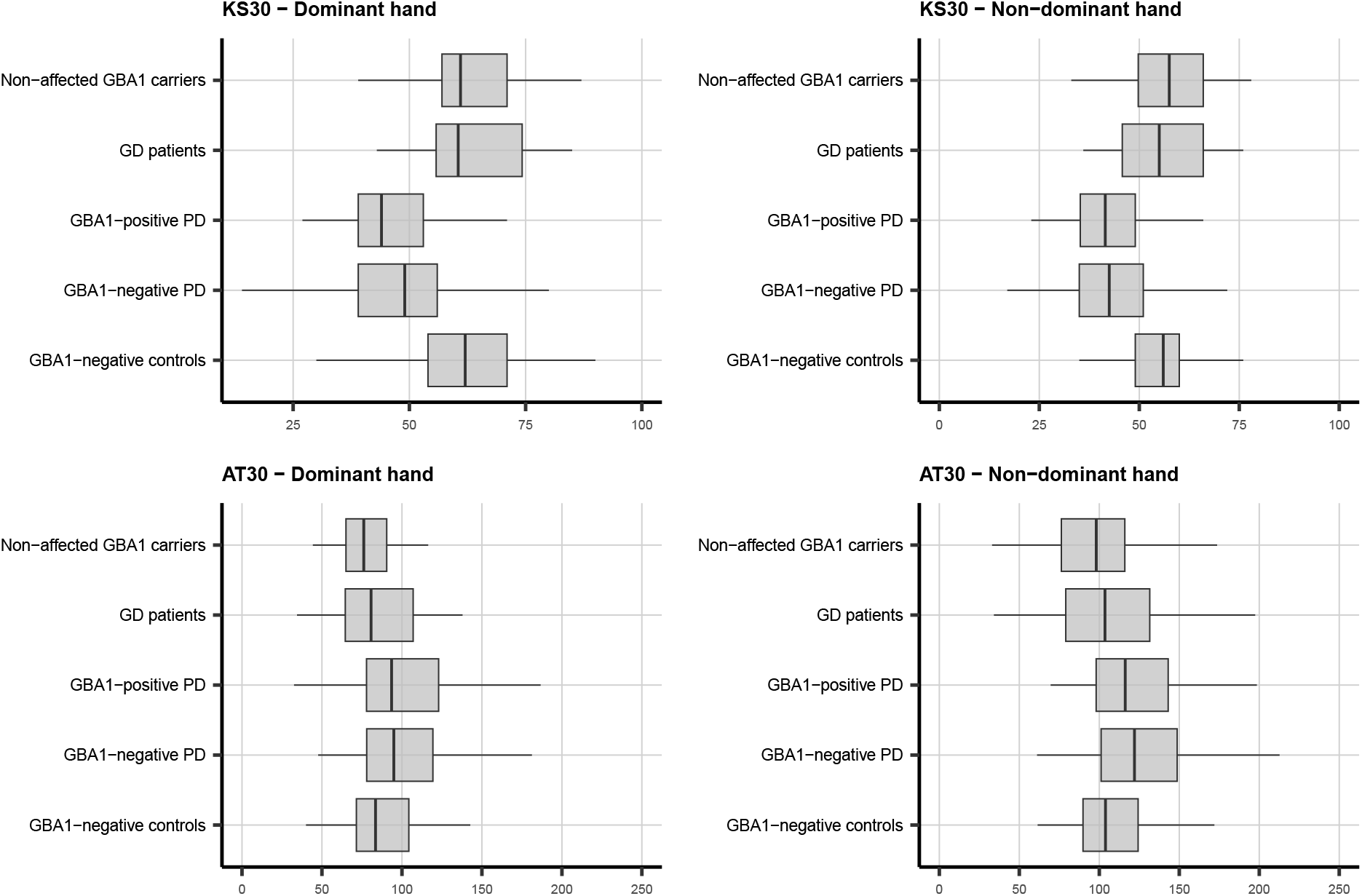
Tap-test data KS30 is reported as number of taps in 30 seconds, AT30 shows the mean dwell time on each key in milliseconds. Data are reported separately for dominant and non-dominant hands. Data are reported as mean (central bar), 25^th^ and 75^th^ percentiles (hinges) and the smallest value at most 1.5 * interquartile range of the hinge (whiskers). KS30: Kinesia Score 30 Seconds, AT30: Akinesia Time 30 seconds.

KS30 for both dominant and non-dominant hands were worse in the two PD groups compared to all the non-PD groups (all p-values < 0.001).

AT30 scores for dominant and non-dominant hands were lower in the two PD groups compared to non-affected *GBA1* carriers and *GBA1*-negative controls (all p-values < 0.01) but were not significantly different from those of GD patients.

KS30 scores were marginally worse in *GBA1*-positive PD patients compared to *GBA1*-negative PD patients for the dominant hand (β = −3.34, p-value = 0.12) and non-dominant hand (β = −3.79, p-value = 0.05).

AT30 score for the non-dominant hand was marginally worse in *GBA1*-positive PD patients compared to *GBA1*-negative PD patients (β = 21.8, p-value = 0.09).

No differences were observed between any of the non-PD groups for KS30 or AT30.

No differences were observed between risk, mild and severe variant carriers among *GBA1*-positive PD and non-affected *GBA1*-carriers.

Results did not change when analysing the RAPSODI cohort separately.

### Cognitive tests

Results of the cognitive tests are reported in supplementary table 4, Figure 3 and supplementary figure 1.

**Figure 3.**
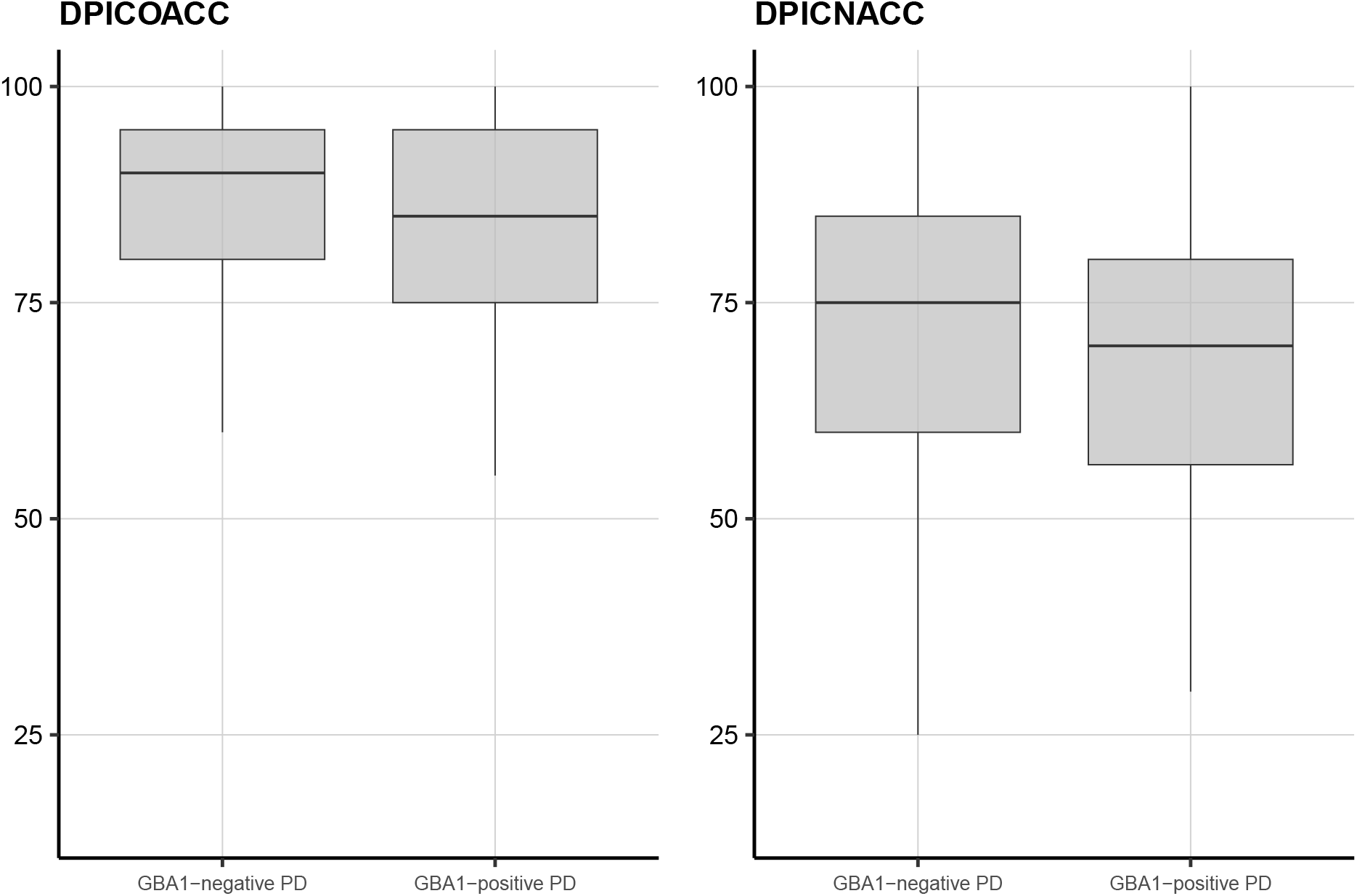
Pattern separation test, differences between *GBA1*-positive and *GBA1*-negative PD Data are reported as mean (central bar), 25^th^ and 75^th^ percentiles (hinges) and the smallest value at most 1.5 * interquartile range of the hinge (whiskers). DPICOACC: Percentage of correct answers recognising original pictures, DPICNACC: Percentage of correct answers recognising new pictures.

The scores of the pictures recognition test (DPICOACC and DPICNACC) and reaction time (SRT, CRT, SPMRT, NWMRT, VIGRT) were worse in the two PD groups compared to the non-PD groups (all p-values < 0.05).

When comparing *GBA1*-positive and *GBA1*-negative PD patients only, *GBA1*-positive PD patients showed a significantly worse performance for DPICOACC, DPICNACC and CRTM (p-values 0.015, 0.039 and 0.0246, respectively – shown in figure 3).

There were no statistically significant differences between the two PD groups for CRTACC, VIGACC, SPMOACC, SPMNACC, NWMOACC, NWMNACC.

Moreover, no significant differences were observed between the non-PD groups for any of the tests.

No differences were observed between risk, mild and severe variant carriers among *GBA1*-positive PD and non-affected *GBA1*-carriers.

## DISCUSSION

In this study, we analysed baseline data from the RAPSODI portal, comparing characteristics between 5 groups: *GBA1*-positive PD patients, *GBA1*-negative PD patients, non-affected *GBA1* carriers, *GBA1*-negative controls, GD patients. We sought further validation of the data by including participants to the PREDICT-PD cohort.

For most of the captured outcomes, both groups of PD patients performed significantly worse compared to people without PD, suggesting that the assessment tools are appropriate for capturing differences between these two populations. Analysis of longitudinal data will clarify whether the assessment is also able to detect subtle changes in currently unaffected individuals that might then develop PD.

We showed a difference in the PD phenotype of *GBA1* carriers compared to non-carriers in UPSIT, tap test and cognitive tests for pattern recognition and reaction time. For some of the other scores, even when not statistically significant, the data suggested a trend toward a worse performance of *GBA1*-positive PD patients compared to *GBA1*-negative PD patients (constipation, anxiety and depression, RBD, working memory).

A previous study similarly showed a worse cognitive profile in 26 *GBA1*-positive PD compared to 39 *GBA1*-negative PD, but no differences in UPSIT(3), and another study showed a more pronounced progression of cognitive dysfunction in 59 *GBA1*-positive PD compared to 684 *GBA1*-negative PD (24). On the other hand, a recent study showed no differences in the cognitive profile in PD patients with or without *GBA1* variants and duration of disease <3.5 years (193 *GBA1*-PD vs 1700 *GBA1*-negative PD)(5). Recent analysis of the large Parkinson’s Progression Markers Initiative (PPMI) cohort showed no difference in olfaction between *GBA1* positive and *GBA1* negative PD patients.

Our findings support the notion that cognition is more affected in *GBA1*-positive PD patients and suggest that olfaction is also worse in *GBA1*-positive PD patients, calling for additional confirmation in independent cohorts.

Of interest is the difference between the two PD groups in the pattern recognition test, which involves visual memory and visuospatial skills, supporting previous evidence that visual functions are more affected in *GBA1*-positive PD(3,25,26).

We did not observe a significantly different age at onset of PD or a different prevalence of males and females, as has been reported in other studies(5).

Moreover, we did not detect a phenotypical effect of *GBA1* variants severity when stratifying them as risk, mild and severe(23). Given the small sample size, the analysis was likely underpowered for this type of analysis.

It remains uncertain as to whether non-affected *GBA1* variant carriers show a higher prevalence of prodromal PD features than the general population.

A previous cohort study from our group showed worse olfaction, cognition and motor signs of PD at baseline, and a steeper progression, in *GBA1* variant carriers compared to non-carrier controls. This cohort had a smaller sample size, and most of the differences between the groups were already present at baseline. A recent study showed no significant deterioration of UPSIT scores in 117 unaffected *GBA1* variants carriers compared to controls(12).

The cross-sectional analysis presented in our paper did not highlight any significant differences between heterozygous and biallelic *GBA1* variant carriers and *GBA1*-negative controls. The longitudinal assessment will clarify whether the two groups show a different rate of progression of prodromal PD symptoms or conversion to PD. Whether this hypothetical difference in prodromal symptoms simply reflects the *GBA1* genotype status or truly represents an early manifestation of PD, will also remain an open question that longitudinal studies will address.

Our studies use an online approach to assess participants. This enables us to reach a larger audience and facilitates participation.

However, this process has limitations. First, there might be a selection bias toward more computer literate individuals, as participants that do not own a computer, or that do not know how to use one, are automatically excluded from the trial. Moreover, most of the assessment is unsupervised, with an intrinsic risk of introducing unreliable observations (participants might ask for help to complete some tasks, there might be connectivity issues hindering the assessment, some instructions on how to carry out the tests might be misunderstood). We addressed these issues by using the median response times in the cognitive tests, a parameter that is less affected by extreme outliers.

Another potential limitation of this study is the selection of *GBA1*-negative controls among relatives (especially partners and spouses) of *GBA1* carriers and PD and GD patients. This has the advantage of including controls that are exposed to similar environmental factors, but the disadvantage of creating a group that is inherently mismatched for sex.

In conclusion, we were able to show a different phenotype in *GBA1* positive PD patients compared to *GBA1* negative PD patients, with the former having worse olfaction and cognitive performance (visual function and reaction time). We did not show any meaningful differences between *GBA1*-negative controls and non-affected *GBA1* carriers.

The analysis of the longitudinal data will provide additional insight into differences in progression between these groups.

## Supporting information

Participant level data

## AKNOWLEDGEMENTS

We would like to thank AH software for their support in designing and maintaining the RAPSODI online portal.

We would like to thank Professor Keith Andrew Wesnes for his support with collecting and interpreting the CogTrack data.

## DATA AVAILABILITY

Participant level data are reported as supplementary material in the file named “participant-level data”.

All other data produced in the present study are available upon reasonable request to the authors.

## AUTHORS ROLES

**Toffoli M** Study conception, bioinformatic & Statistical Analysis, Writing of the first draft

**Chohan H** Data Collection, Manuscript Review

**Mullin S** Study conception, Manuscript Review

**Jesuthasan A**Data Collection, Manuscript Review Yalkic S Data Collection, Manuscript Review

**Koletsi S** Data Collection, Manuscript Review

**Menozzi E** Data Collection, Manuscript Review

**Rahall S** Data Collection, Manuscript Review

**Limbachiya N** Data Collection, Manuscript Review

**Loefflad N** Data Collection, Manuscript Review

**Higgins A** Data Collection, Manuscript Review

**Bestwick J** Data Collection, Manuscript Review

**Lucas-Del-Pozo S** Data Collection, Manuscript Review

**Fierli F** Data Collection, Manuscript Review

**Farbos A** Genetic analysis

**Mezabrovschi R** Data Collection, Manuscript Review

**Lee-Yin C** Data Collection, Manuscript Review

**Schrag A** Study conception, Manuscript Review

**Moreno-Martinez D** Data Collection, Manuscript Review

**Hughes D** Study conception, Data Collection, Manuscript Review

**Noyce A** Study conception, data collection, Manuscript Review

**Colclough K** Genetic analysis, Manuscript Review

**Jeffries AR** Genetic analysis, Manuscript Review

**Proukakis C** Study conception, Manuscript Review

**Schapira AHV** Study conception and supervision, Manuscript Review

## ETHICAL COMPLIANCE STATEMENT

All participants gave informed consent to be included in the study. The work was approved by the London – Queen Square Research Ethics Committee (REC reference: 15/LO/1155). We confirm that we have read the Journal’s position on issues involved in ethical publication and affirm that this work is consistent with those guidelines.

Ethical approval for the PREDICT-PD study was grant by Central London Research Ethics Committee 3 (reference number 10/H0716/85).

