## Supplemental figures and tables for "Phenotypic effect of *GBA1* variants in individuals with and without Parkinson disease: the RAPSODI study"

### Supplementary

#### Supplementary Figure 1 – Results of the cognitive tests

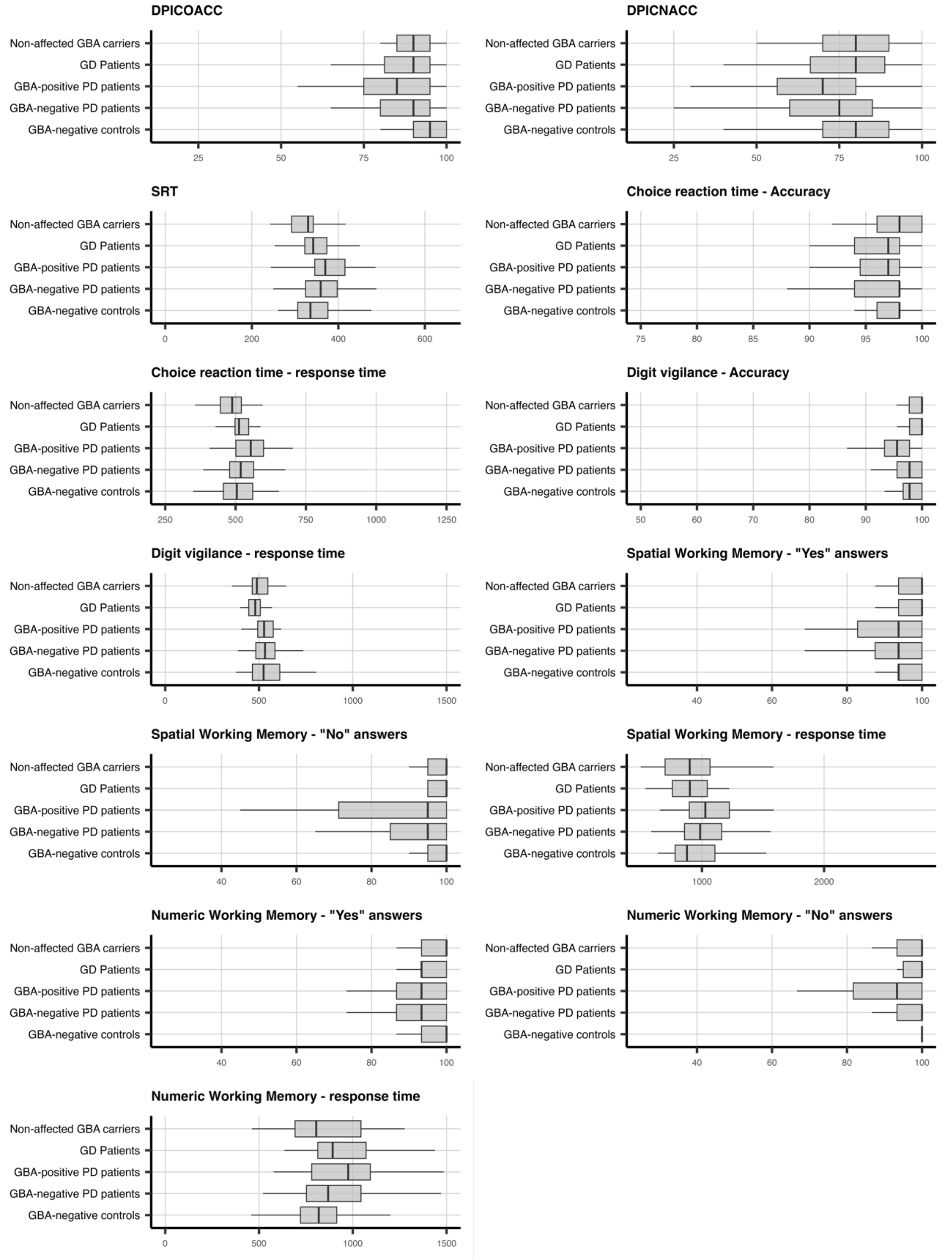

For a description of each score, please see Table 1.
Data are reported as mean (central bar), $25^{th}$ and $75^{th}$ percentiles (hinges) and the smallest value at most 1.5 * interquartile range of the hinge (whiskers).

Supplementary Table 1 – *GBA2* genotype of participants

| ***GBA1*-positive PD patients** | **Number of participants** |
|---|---|
| **E365K / WT** | 21 |
| **T408M / WT** | 7 |
| **N409S / WT** | 5 |
| **L483P / WT** | 3 |
| **R301H / WT** | 2 |
| **A495P Val499= L483P / WT** | 1 |
| **c.413delC / WT** | 1 |
| **G241R / WT** | 1 |
| **K13R / WT** | 1 |
| **R502C / WT** | 1 |
| **R535H / WT** | 1 |
| **T408M + W432ter / WT** | 1 |
| **V433L / WT** | 1 |
| **WT / A495P Val499= L483P** | 1 |
| **GD Patients** | |
| **N409S / L483P** | 6 |
| **L483P / N409S** | 4 |
| **N409S / N409S** | 4 |
| **N409S / A495P Val499= L483P** | 3 |
| **N409S / IVS9+1** | 2 |
| **D419N / N409S** | 1 |
| **N409S / D354H** | 1 |
| **N409S / G241R** | 1 |
| **N409S / G289V** | 1 |
| **N409S / L144R** | 1 |
| **N409S / L519P** | 1 |
| **N409S / P221T** | 1 |
| **N409S / R398ter** | 1 |
| **Non-affected *GBA1* carriers** | |
| **N409S / WT** | 24 |
| **E365K / WT** | 27 |
| **T408M / WT** | 13 |
| **L483P / WT** | 6 |
| **R502C / WT** | 3 |
| **A495P Val499= L483P / WT** | 2 |
| **84GG / WT** | 2 |

| | |
|---|---|
| **L519P / WT** | 2 |
| **V433L / WT** | 2 |
| **T408M / T408M** | 1 |
| **E365K / E365K** | 1 |
| **G154R / WT** | 1 |
| **IVS6-2 / WT** | 1 |
| **IVS9+1 / WT** | 1 |
| **R209P / WT** | 1 |
| **R535H / WT** | 1 |
| **S212Ter / WT** | 1 |
| **T270I / WT** | 1 |

#### Supplementary Table 2 – Clinical questionnaires and UPSIT

| Do you suffer from hard stools | | | | | | |
|---|---|---|---|---|---|---|
| status | No | Sometimes | Yes | No | Sometimes | Yes |
| *GBA1*-negative controls | 82 | 48 | 9 | 59.0% | 34.5% | 6.5% |
| *GBA1*-negative PD patients | 80 | 88 | 14 | 44.0% | 48.4% | 7.7% |
| *GBA1*-positive PD patients | 26 | 16 | 11 | 49.1% | 30.2% | 20.8% |
| GD Patients | 17 | 13 | 3 | 51.5% | 39.4% | 9.1% |
| Non-affected *GBA1* carriers | 55 | 29 | 1 | 64.7% | 34.1% | 1.2% |

| Does opening your bowels require a lot of effort | | | | | | |
|---|---|---|---|---|---|---|
| status | No | Sometimes | Yes | No | Sometimes | Yes |
| *GBA1*-negative controls | 89 | 46 | 4 | 64.0% | 33.1% | 2.9% |
| *GBA1*-negative PD patients | 62 | 99 | 21 | 34.1% | 54.4% | 11.5% |
| *GBA1*-positive PD patients | 15 | 26 | 12 | 28.3% | 49.1% | 22.6% |
| GD Patients | 23 | 7 | 3 | 69.7% | 21.2% | 9.1% |
| Non-affected *GBA1* carriers | 60 | 23 | 2 | 70.6% | 27.1% | 2.4% |

| Do you ever use laxatives | | | | | | |
|---|---|---|---|---|---|---|
| status | No | Sometimes | Yes | No | Sometimes | Yes |
| *GBA1*-negative controls | 126 | 8 | 10 | 87.5% | 5.6% | 6.9% |
| *GBA1*-negative PD patients | 114 | 34 | 34 | 62.6% | 18.7% | 18.7% |
| *GBA1*-positive PD patients | 31 | 9 | 13 | 58.5% | 17.0% | 24.5% |
| GD Patients | 26 | 4 | 3 | 78.8% | 12.1% | 9.1% |
| Non-affected *GBA1* carriers | 83 | 4 | 5 | 90.2% | 4.3% | 5.4% |

| MDS-UPDRS2 | | | | |
|---|---|---|---|---|
| status | Count | Mean | Sd | Median |

| | | | | |
|---|---|---|---|---|
| *GBA1*-negative PD patients | 181 | 10 | 7.1 | 9 |
| *GBA1*-negative controls | 100 | 0.9 | 2.5 | 0 |
| *GBA1*-positive PD patients | 52 | 11.5 | 7 | 11 |
| GD Patients | 25 | 2.1 | 2.7 | 1 |
| Non-affected *GBA1* carriers | 43 | 1 | 1.6 | 0 |

| HADS - Anxiety | | | | | | |
|---|---|---|---|---|---|---|
| | Abnormal | Borderline | Normal | Abnormal | Borderline | Normal |
| *GBA1*-negative controls | 5 | 20 | 110 | 3.7% | 14.8% | 81.5% |
| *GBA1*-negative PD patients | 22 | 28 | 116 | 13.3% | 16.9% | 69.9% |
| *GBA1*-positive PD patients | 7 | 13 | 31 | 13.7% | 25.5% | 60.8% |
| GD Patients | 3 | 0 | 28 | 9.7% | 0.0% | 90.3% |
| Non-affected *GBA1* carriers | 6 | 7 | 77 | 6.7% | 7.8% | 85.6% |

| HADS - Depression | | | | | | |
|---|---|---|---|---|---|---|
| | Abnormal | Borderline | Normal | Abnormal | Borderline | Normal |
| *GBA1*-negative controls | 1 | 4 | 134 | 0.7% | 2.9% | 96.4% |
| *GBA1*-negative PD patients | 7 | 18 | 142 | 4.2% | 10.8% | 85.0% |
| *GBA1*-positive PD patients | 4 | 5 | 34 | 9.3% | 11.6% | 79.1% |
| GD Patients | 1 | 2 | 28 | 3.2% | 6.5% | 90.3% |
| Non-affected *GBA1* carriers | 0 | 1 | 91 | 0.0% | 1.1% | 98.9% |

| RBDsq | | | | |
|---|---|---|---|---|
| | No | Yes | No | Yes |
| *GBA1*-negative controls | 130 | 14 | 90.3% | 9.7% |
| *GBA1*-negative PD patients | 125 | 60 | 67.6% | 32.4% |
| *GBA1*-positive PD patients | 27 | 24 | 52.9% | 47.1% |
| GD Patients | 29 | 5 | 85.3% | 14.7% |
| Non-affected *GBA1* carriers | 90 | 6 | 93.8% | 6.2% |

| UPSIT | | | | | | | | | | |
|---|---|---|---|---|---|---|---|---|---|---|
| | Total anosmia | Severe microsmia | Moderate microsmia | Mild microsmia | Normosmia | Total anosmia | Severe microsmia | Moderate microsmia | Mild microsmia | Normosmia |
| GBA-negative controls | 1 | 8 | 16 | 51 | 47 | 0.8% | 6.5% | 13.0% | 41.5% | 38.2% |
| GBA-negative PD patients | 64 | 48 | 29 | 18 | 10 | 37.9% | 28.4% | 17.2% | 10.7% | 5.9% |
| GBA-positive PD patients | 23 | 17 | 5 | 0 | 2 | 48.9% | 36.2% | 10.6% | 0.0% | 4.3% |
| GD Patients | 2 | 1 | 3 | 13 | 9 | 7.1% | 3.6% | 10.7% | 46.4% | 32.1% |
| Non-affected GBA carriers | 3 | 6 | 12 | 32 | 27 | 3.8% | 7.5% | 15.0% | 40.0% | 33.8% |

Supplementary Table 3 – Tap test

| KS30 - Dominant | | | | |
|---|---|---|---|---|
| status | Count | Mean | Sd | Median |
| *GBA-negative PD patients* | 179 | 48.9 | 13.1 | 49.0 |
| *GBA-negative controls* | 136 | 61.9 | 13.1 | 62.0 |
| *GBA-positive PD patients* | 49 | 47.1 | 12.9 | 44.0 |
| GD Patients | 24 | 63.6 | 12.3 | 60.5 |
| Non-affected GBA carriers | 77 | 62.5 | 12.6 | 61.0 |

| KS30 - Non-dominant | | | | |
|---|---|---|---|---|
| status | Count | Mean | Sd | Median |
| *GBA-negative PD patients* | 176 | 44.0 | 12.4 | 42.5 |
| *GBA-negative controls* | 136 | 55.3 | 10.5 | 56.0 |
| *GBA-positive PD patients* | 50 | 41.4 | 11.6 | 41.5 |
| GD Patients | 24 | 55.6 | 12.8 | 55.0 |
| Non-affected GBA carriers | 76 | 57.3 | 10.2 | 57.5 |

| AT30 - Dominant | | | | |
|---|---|---|---|---|
| status | Count | Mean | Sd | Median |
| *GBA-negative PD patients* | 179 | 104.0 | 39.2 | 95.2 |
| *GBA-negative controls* | 136 | 89.6 | 28.0 | 83.5 |
| *GBA-positive PD patients* | 49 | 101.9 | 34.4 | 93.5 |
| GD Patients | 24 | 92.9 | 44.2 | 84.4 |
| Non-affected GBA carriers | 77 | 85.2 | 40.6 | 76.3 |

| AT30 - Non-dominant | | | | |
|---|---|---|---|---|
| status | Count | Mean | Sd | Median |
| *GBA-negative PD patients* | 176 | 131.8 | 46.9 | 122.1 |
| *GBA-negative controls* | 136 | 111.3 | 31.1 | 103.9 |
| *GBA-positive PD patients* | 50 | 148.5 | 142.6 | 117.3 |
| GD Patients | 24 | 116.2 | 51.2 | 103.6 |
| Non-affected GBA carriers | 76 | 101.3 | 34.8 | 100.6 |

Supplementary Table 4 – Cognitive tests

| **DPICOACC** | | | | |
|---|---|---|---|---|
| **status** | **Count** | **Mean** | **Sd** | **Median** |
| *GBA1*-negative PD patients | 176 | 87.3 | 10.2 | 90 |
| *GBA1*-negative controls | 83 | 92.3 | 7.5 | 95 |
| *GBA1*-positive PD patients | 34 | 82.5 | 15.7 | 85 |
| GD Patients | 26 | 88.5 | 11.4 | 90 |
| Non-affected GBA carriers | 37 | 88.8 | 10.7 | 90 |

| **DPICNACC** | | | | |
|---|---|---|---|---|
| **status** | **Count** | **Mean** | **Sd** | **Median** |
| *GBA1*-negative PD patients | 176 | 73.0 | 15.3 | 75 |
| *GBA1*-negative controls | 83 | 78.4 | 15.1 | 80 |
| *GBA1*-positive PD patients | 34 | 68.5 | 18.2 | 70 |
| GD Patients | 26 | 76.3 | 16.5 | 80 |
| Non-affected GBA carriers | 37 | 79.5 | 16.1 | 80 |

| **SRT** | | | | |
|---|---|---|---|---|
| **status** | **Count** | **Mean** | **Sd** | **Median** |
| *GBA1*-negative PD patients | 176 | 380.2 | 102.8 | 361.4 |
| *GBA1*-negative controls | 83 | 346.1 | 56.6 | 335.5 |
| *GBA1*-positive PD patients | 34 | 389.6 | 82.4 | 369.8 |
| GD Patients | 26 | 349.7 | 48.7 | 341.9 |
| Non-affected GBA carriers | 37 | 330.7 | 53.5 | 330.1 |

| **CRTACC** | | | | |
|---|---|---|---|---|
| **status** | **Count** | **Mean** | **Sd** | **Median** |
| *GBA1*-negative PD patients | 176 | 96.2 | 3.8 | 98 |
| *GBA1*-negative controls | 83 | 97.1 | 2.4 | 98 |
| *GBA1*-positive PD patients | 34 | 95.5 | 4.5 | 97 |
| GD Patients | 26 | 96.3 | 2.8 | 97 |
| Non-affected GBA carriers | 37 | 96.4 | 4.2 | 98 |

| **CRT** | | | | |
|---|---|---|---|---|
| **status** | **Count** | **Mean** | **Sd** | **Median** |
| *GBA1*-negative PD patients | 176 | 543.7 | 121.9 | 518.7 |
| *GBA1*-negative controls | 83 | 512.4 | 74.0 | 504.5 |
| *GBA1*-positive PD patients | 34 | 572.6 | 120.2 | 554.1 |
| GD Patients | 26 | 514.6 | 55.8 | 512.3 |

| | | | | |
|---|---|---|---|---|
| Non-affected GBA carriers | 37 | 489.5 | 80.8 | 487.9 |

| **VIGACC** | | | | |
|---|---|---|---|---|
| **status** | **Count** | **Mean** | **Sd** | **Median** |
| *GBA1*-negative PD patients | 176 | 95.3 | 10.5 | 97.8 |
| *GBA1*-negative controls | 83 | 96.9 | 5.1 | 97.8 |
| *GBA1*-positive PD patients | 34 | 93.1 | 12.0 | 95.6 |
| GD Patients | 26 | 97.4 | 5.1 | 100.0 |
| Non-affected GBA carriers | 37 | 97.7 | 3.5 | 100.0 |

| **VIGRT** | | | | |
|---|---|---|---|---|
| **status** | **Count** | **Mean** | **Sd** | **Median** |
| *GBA1*-negative PD patients | 176 | 596.7 | 255.1 | 534.6 |
| *GBA1*-negative controls | 83 | 563.7 | 134.2 | 524.3 |
| *GBA1*-positive PD patients | 34 | 614.7 | 306.2 | 534.0 |
| GD Patients | 26 | 485.8 | 53.5 | 479.9 |
| Non-affected GBA carriers | 37 | 511.4 | 91.9 | 488.6 |

| **SPMOACC** | | | | |
|---|---|---|---|---|
| **status** | **Count** | **Mean** | **Sd** | **Median** |
| *GBA1*-negative PD patients | 175 | 90.0 | 14.7 | 93.8 |
| *GBA1*-negative controls | 83 | 91.9 | 13.2 | 93.8 |
| *GBA1*-positive PD patients | 34 | 87.9 | 16.6 | 93.8 |
| GD Patients | 26 | 93.8 | 10.8 | 100.0 |
| Non-affected GBA carriers | 37 | 94.4 | 7.0 | 100.0 |

| **SPMNACC** | | | | |
|---|---|---|---|---|
| **status** | **Count** | **Mean** | **Sd** | **Median** |
| *GBA1*-negative PD patients | 175 | 88.2 | 18.4 | 95.0 |
| *GBA1*-negative controls | 83 | 93.7 | 12.4 | 100.0 |
| *GBA1*-positive PD patients | 34 | 85.9 | 17.8 | 95.0 |
| GD Patients | 26 | 95.2 | 10.4 | 100.0 |
| Non-affected GBA carriers | 37 | 96.4 | 6.6 | 100.0 |

| **SPMRT** | | | | |
|---|---|---|---|---|
| **status** | **Count** | **Mean** | **Sd** | **Median** |
| *GBA1*-negative PD patients | 175 | 1055.6 | 385.3 | 985.0 |
| *GBA1*-negative controls | 83 | 958.0 | 280.5 | 877.1 |
| *GBA1*-positive PD patients | 34 | 1092.9 | 360.7 | 1027.8 |

| | | | | |
|---|---|---|---|---|
| GD Patients | 26 | 917.8 | 248.6 | 900.8 |
| Non-affected GBA carriers | 37 | 933.9 | 447.6 | 899.5 |

| **NWMOACC** | | | | |
|---|---|---|---|---|
| **status** | **Count** | **Mean** | **Sd** | **Median** |
| *GBA1*-negative PD patients | 176 | 90.1 | 14.3 | 93.3 |
| *GBA1*-negative controls | 83 | 94.5 | 7.4 | 100.0 |
| *GBA1*-positive PD patients | 34 | 89.0 | 12.6 | 93.3 |
| GD Patients | 26 | 93.8 | 6.2 | 93.3 |
| Non-affected GBA carriers | 37 | 96.4 | 5.8 | 100.0 |

| **NWMNACC** | | | | |
|---|---|---|---|---|
| **status** | **Count** | **Mean** | **Sd** | **Median** |
| *GBA1*-negative PD patients | 176 | 90.1 | 14.3 | 93.3 |
| *GBA1*-negative controls | 83 | 94.5 | 7.4 | 100.0 |
| *GBA1*-positive PD patients | 34 | 89.0 | 12.6 | 93.3 |
| GD Patients | 26 | 93.8 | 6.2 | 93.3 |
| Non-affected GBA carriers | 37 | 96.4 | 5.8 | 100.0 |

| **NWMRT** | | | | |
|---|---|---|---|---|
| **status** | **Count** | **Mean** | **Sd** | **Median** |
| *GBA1*-negative PD patients | 176 | 881.9 | 203.8 | 851.5 |
| *GBA1*-negative controls | 83 | 807.6 | 163.6 | 787.0 |
| *GBA1*-positive PD patients | 34 | 905.5 | 175.3 | 942.7 |
| GD Patients | 26 | 878.2 | 198.1 | 841.0 |
| Non-affected GBA carriers | 37 | 811.7 | 207.1 | 773.0 |
